# The genetic landscape of kynurenine predicts neurovascular pathology and disrupted white matter integrity in patients with mood disorders

**DOI:** 10.1101/2024.12.11.24317786

**Authors:** Beatrice Bravi, Lidia Fortanyer-Uyà, Marco Paolini, Stefano Comai, Sara Poletti, Lorenzi Cristina, Sara Spadini, Alessandro Serretti, Cristina Colombo, Raffaella Zanardi, Francesco Benedetti

## Abstract

Low-grade systemic inflammation is linked to cardiometabolic diseases and increased cardiovascular risk. Patients with mood disorders, such as Major Depressive Disorder (MDD) and Bipolar Disorder (BD), also show elevated cardiovascular risk and inflammatory markers, suggesting shared biological pathways between mood and cardiometabolic conditions. The kynurenine (KYN) pathway, activated by inflammatory cytokines and involved in neurotransmitter systems linked to mood, provides a promising area to explore inflammatory-related genetic overlaps in these disorders, with increasing interest in the SH2B3 rs3184504 SNP. Imaging markers like white matter hyperintensities (WMHs) and white matter (WM) microstructure alterations are associated with mood and cardiovascular disorders.

This study aimed to investigate the genetic load link to KYN levels, such as KYN polygenic risk score (PRS) and its effect on white matter hyperintensities (WMHs), outcomes of presumed vascular suffering, and WM microstructure in a sample of 95 MDD and 80 BD patients.

Higher PRS for KYN was associated with increased circulating KYN levels and KYN/TRP ratio. KYN PRS predicted the presence of WMHs. The SH2B3 rs3184504 T variant was associated with increased PRS for KYN and a higher number of WMHs. KYN levels and KYN/TRP ratio were not associated with WMHs, while KYN PRS positively correlated with higher axial (AD) and mean diffusivity (MD), with a nominal significance for radial diffusivity (RD).

The findings support a genetic contribution to elevated KYN and WM integrity alterations in mood disorders. PRS for KYN indicates a potential predisposition to inflammatory and vascular dysregulation, and SH2B3 rs3184504 may modulate this risk.

## Introduction

Low-grade systemic inflammation is associated with the development and progression of cardiometabolic medical conditions in the general population, substantially increasing the risk for cardiovascular disease; in turn, patients with cardiometabolic conditions have raised markers of systemic inflammation (Furman et al., 2019; Mainous III et al., 2024; Pearson et al., 2003). Patients with mood disorders are at higher risk for the subsequent onset of several medical conditions, including cardiovascular diseases (Momen et al., 2020), with lifetime disease trajectories suggesting a direct link between psychiatric disorders and cardiovascular diseases, with or without mediating medical conditions (Shen et al., 2023). Low-grade systemic inflammation could contribute to this link. Circulating inflammatory biomarkers are markedly raised both, in Major Depressive Disorder (MDD) and in Bipolar Disorder (BD), in the absence of known underpinning medical conditions (Poletti et al., 2024), and abnormally high immune-inflammatory setpoints are currently regarded as pathophysiological mechanisms and possible targets for treatment in mood disorders (Benedetti et al., 2022; Branchi et al., 2021). The study of dysregulated homeostatic pathways also suggests a clustering of immunometabolic biological dysregulations in mood disorders, underpinning both, depressive psychopathology and comorbid medical conditions (Milaneschi et al., 2020), and fostering a worse neuroprogression.

Genetic factors could contribute to this overlap of higher pro-inflammatory and cardiometabolic altered phenotypes in mood disorders, since meta-analyses of genome-wide and candidate gene association studies suggested shared biological mechanisms of mood disorders and cardiometabolic diseases, with pleiotropic genes and biological pathways common to the two categories of disease (Amare et al., 2017). Following this perspective, the kynurenine pathway appears a good candidate to study the genetic overlap between inflammation and cardiometabolic risk in mood disorders.

Patients with mood disorders show a higher activation of indoleamine-2,3-dioxygenase (IDO), which is activated by proinflammatory cytokines, and converts tryptophan (TRP) into kynurenine (KYN) (Leonard, 2001). TRP breakdown can take place in astrocytes, microglia, and neurons, but about 60% of brain KYN is transported across the blood-brain barrier (BBB) from the periphery, and is then circulated into the brain via volume transmission (Fuxe et al., 2013). IDO activation targets the same neurotransmitter systems involved in mood disorders, by decreasing the availability of TRP for serotonin synthesis, and by directly acting on glutamatergic transmission at the NMDAR level, via the KYN metabolites kynurenic and quinolinic acid (Dantzer et al., 2008). The activation of the KYN pathway can thus be considered as a link between low-grade inflammation and abnormal neurotransmission in mood disorders. Indeed, the KP is regulated by, and in turn regulates multiple other physiological systems that are commonly disrupted in psychiatric disorders, including endocrine, metabolic, and hormonal systems (Savitz, 2020).

Genetic factors increasing circulating levels of KYN proportionally increase the risk of depression (Zong et al., 2024). Beyond neurotransmitters, studies in the general population suggest that the genetic architecture of plasma kynurenine includes cardiometabolic disease mechanisms (Bagheri et al., 2021). In particular, among several single nucleotide polymorphisms (SNPs) associated with circulating levels of KYN, the SH2B3 rs3184504 C>T nonsynonymous SNP emerged as a genetic determinant of both, plasma KYN levels, and cardiovascular diseases, with the missense T variant associated with a range of conditions including hypertension, atherosclerosis and myocardial infarction at phenome-wide association testing (Bagheri et al., 2021). Part of the Src homology 2-B adapter family, the SH2B3 gene in humans codes for the lymphocyte adaptor protein LNK, which is widely expressed in all hematopoietic lineages and some non-hematopoietic cells, particularly endothelial cells, to act as a negative key regulator of cytokine receptor-mediated signaling and cell proliferation (Devallière and Charreau, 2011; Morris et al., 2021). The missense T variant results in the substitution of TRP for arginine at amino acid 262, causing a loss of LNK function, and resulting in hypersensitivity to cytokines, enhanced JAK/STAT signaling and effector functions in CD8+ T cells, and, after stimulation, a 3- to 5-fold higher production of proinflammatory cytokines in homozygous TT human individuals (Alexander et al., 2022; Watson et al., 2024; Zhernakova et al., 2010): animal models also suggest that this phenotype might improve survival and morbidity with less organ damage and earlier bacterial clearance in sepsis, at the price of a higher risk of hypertension, cardiovascular disease, and medical conditions associated with altered immunity, as documented in humans (Allenspach et al., 2021).

In previous studies, we associated the activation of IDO and the breakdown ratio of TRP to kynurenine with immune-inflammatory markers, and with detrimental effects in the brain of patients with mood disorders, including lower cortical thickness and disrupted white matter (WM) microstructure. In light of the above, it is tempting to hypothesize that genetic factors affecting the KYN pathway could influence neurovascular risk and white matter integrity in patients affected by mood disorders.

The purpose of the present study was then to examine the effect, in a sample of 175 patients with BD or MDD, of a polygenic risk score (PRS) for KYN, derived from a large GWAS study (Chen et al., 2023), and of the rs3184504 C>T gene variant, on: (i) circulating levels of KYN and TRP breakdown ratio; (ii) the number of white matter hyperintensities (WMHs) of presumed vascular origin, which are one of the imaging markers of microvascular pathology and neurovascular inflammation (Chen et al., 2021; Huang et al., 2022); and (iii) in vivo measures of WM microstructure at magnetic resonance diffusion-tensor imaging (DTI).

## 2. Experimental procedures

### 2.1 Sample/Participants

The sample included 95 MDD and 80 BD patients (DSM-5) consecutively admitted to the IRCCS San Raffaele Turro in Milan, Italy.

Inclusion criteria were: age between 18-65 years old; willingness to participate; absence of other diagnoses on Axis I, pregnancy, history of epilepsy, major medical and neurological disorders; intellectual disabilities; absence of a history of drug or alcohol dependency; absence of inflammation-related diseases or symptoms, including fever and infections; no autoimmune diseases; no uncontrolled systemic, metabolic or other significant somatic disorders known to affect mood; somatic medications known to affect mood or the immune system, such as corticosteroids, non-steroid anti-inflammatory drugs and statins.

Clinical data were collected through structured psychiatric interviews. Fasting blood samples were taken in the morning (between 7:00 to 9:00 a.m.). Patients’ ongoing drug treatments were calculated as equivalent doses of chlorpromazine, imipramine, and lorazepam.

Written informed consent was obtained after a complete description of the study to the subjects. The local ethical committee approved all the research activities (IRCCS Ospedale San Raffaele) and complied with the Helsinki Declaration of 1975, as revised in 2013.

### 2.2 TRP and KYN plasma levels

Plasma levels of TRP and KYN were assessed using a standard methodology in our laboratory (Comai et al., 2022; Simonato et al., 2021). Blood was collected by venipuncture in Vacutainer tubes containing EDTA in the morning after an overnight fast, centrifuged at 2000 ×g for 15 min at 4 °C, and the plasma was divided into small aliquots and stored at -80 °C. Analysis of TRP and KYN was performed using a high-performance liquid chromatography (HPLC) system coupled with a fluorometric detector for TRP (excitation at 285 nm, emission at 345 nm) and a UV-Vis detector for KYN (absorbance at 360 nm). The mobile phase consisted of 10% acetonitrile/90% phosphate buffer 0.004 M at pH 3.5. Isocratic elution at 0.8 mL/min along a Synergi Fusion-RP 80A column (4 μm; 250 × 4.6 mm; Phenomenex, Aschaffenburg, Germany) was performed for separation. The ratio KYN/TRP*1000 was calculated as a proxy of IDO activity.

### 2.3 Genotyping, quality control, and imputation

Genotyping, quality control (QC), and imputation of genetic data were conducted within a larger cohort of 192 MDD patients and 265 BD patients, of which the current sample represents a subset. Blood samples from participants were genotyped using the Infinium PsychArray 24 BeadChip (Illumina, Inc., San Diego), which is a cost-effective, high-density microarray developed in accordance with the Psychiatric Genomics Consortium for large-scale genetic studies focused on psychiatric predispositions and risks. QC of the genotyped data were made according to Anderson et al. (2010) (Anderson et al., 2010) guidelines and was implemented using PLINK 1.9 (Purcell et al., 2007). QC consisted of two principal stages: individual-level QC and variant-level QC. Individuals were excluded if they exhibited a mismatching genotyped sex, a genotype call rate below 95%, and outlying autosomal heterozygosity (Fhet > ±0.2). Genetic variants were removed if their call rate was lower than 98%, if their minor allele frequency (MAF) was lower than 1%, or if they showed significant deviation from Hardy-Weinberg equilibrium (p < 10-8). Additionally, relatedness and population structure were assessed, so that individuals with a recent shared ancestry between third- and second-degree relatives (i.e., identity by descent [IBD] > 0.1875) or those whose genotype distributions deviated more than 5 standard deviations from the mean of the first two principal components were excluded. After QC, genotype frequencies of SH2B3 rs3184504 were extracted. Finally, genotype imputation was performed using the Michigan Imputation Server with Minimac4, version 1.7.1 (https://imputationserver.sph.umich.edu), selecting the Haplotype Reference Consortium panel (HRC r1.1 2016) and the Eagle (version 2.4) phasing algorithm. The resulting post-imputation files contained approximately 14 million genetic variants.

### 2.4 Polygenic risk scores calculation

PRSs for KYN were calculated as the sum of effect alleles at each SNP position, weighted by their effect sizes derived from the largest genome-wide association studies (GWAS) available at the time of the analysis (Chen et al., 2023). Specifically, PRS for each individual were computed using the score function of PLINK 1.9

All KYN weights were estimated using LDpred2-auto, an R-based tool that allows the adjustment of the posterior mean causal effect sizes for each variant included in the GWAS, based solely on the original GWAS’s summary statistics and an external linkage disequilibrium (LD) reference panel (Prive et al., 2021). Remarkably, all variants included in the GWAS’s summary statistics underwent a QC procedure, excluding markers whose standard deviation exceeded the one derived from the reference panel (Prive et al., 2022).

### 2.5 MRI acquisition

All MRI acquisitions were performed at the C.E.R.M.A.C. (Center for Excellence in Magnetic Resonance Imaging in the High Range) of the San Raffaele Hospital in Milan. Philips Ingenia CX (Philips, Netherlands) 3T scan with 32-channel standard quadrature SENSE head coil was used. The T1-weighted MPRAGE and coronal fluid-attenuated inversion recovery (FLAIR) sequences were acquired. T1-w sequence had the following parameters: TR = 8.00 ms, TE = 3.7 ms, FOV = 256 mm, matrix = 256×256, and an in-plane resolution of 1×1 mm, resulting in 182 transversal slices with a thickness of 1 mm. Coronal fluid-attenuated inversion recovery (FLAIR) images. As part of the quality control procedure, T1- and T2-weighted FLAIR images were visually inspected.

The parameters for DWI acquisitions were as follows: TR/TE=5900/78 ms, FoV (mm) 240 (ap), 129 (fh), 232 (rl); acquisition matrix 2.14×2.73×2.30; 56 contiguous, 2.3 mm thick axial slices reconstructed with in-plane pixel size 1.88×1.88×2.30 mm; SENSE acceleration factor=2; 1 b0 and 40 non-collinear directions of diffusion gradients; b value=1000 s/mm2. Fat saturation was performed to avoid chemical shift artefacts.

### 2.6 Imaging processing

Lesions were segmented by the lesion growth algorithm (Schmidt et al., 2012) as implemented in the Lesion Segmentation Tool (LST) toolbox version 3.0.0 (www.statistical-modelling.de/lst.html) for SPM. The algorithm first segments the T1-weighted images into the three primary tissue classes: cerebrospinal fluid (CSF), gray matter (GM), and white matter (WM). This segmentation is then integrated with the coregistered FLAIR image intensities to generate lesion belief maps. These maps are thresholded using a predefined initial value (κ) to produce an initial binary lesion map, which is then expanded by including voxels that exhibit hyperintensity on the FLAIR images. The final output is a lesion probability map. For each subject, the number of lesions was quantified and used for further analyses.

DTI analysis and tensor calculations were performed using the Oxford Center for Functional Magnetic Resonance Imaging of the Brain Software Library (FSL 6.0; www.fmrib.ox.ac.uk/fsl/index.html) (Smith et al., 2004).

The FSL brain extraction tool (BET), which provides a brain mask for each subject, was first used to remove non-brain tissue. Then, the FSL eddy tool was used, which processes a Gaussian process to predict undistorted data to which the actual volumes can be fitted (Andersson and Sotiropoulos, 2015). It can estimate and correct volume-to-volume motion and off-resonance fields, signal dropout due to motion during diffusion encoding, and intra-volume motion (Andersson et al., 2018). Least squares fits were then performed to estimate the fractional anisotropy (FA), eigenvector and eigenvalue maps. Mean diffusivity (MD) was defined as the mean of all three eigenvalues (λ 1 + λ 2 + λ 3)/3, radial diffusivity (RD) as the mean of the second and third eigenvalues (λ 2 + λ 3)/2, and axial diffusivity (AD) as the main diffusion eigenvalue (λ 1). Next, the volumes of all subjects were skeletonised and transformed into a common space as used in Tract-Based Spatial Statistics (TBSS) (Smith et al., 2006). Briefly, all volumes were non-linearly warped to the FMRIB58_FA template provided with FSL (http://www.fmrib.ox.ac.uk/fsl/tbss/FMRIB58_FA.html) and normalised to the Montreal Neurological Institute space using local deformation procedures performed by FMRIB’s Non-Linear Image Registration Tool (FNIRT). A mean FA image of all subjects was then calculated and thinned to create a mean FA skeleton representing the centres of all common tracts. To reduce the likelihood of partial volumes at the boundaries between tissue classes, we applied a threshold (FA > 0.20) and binarised the mean FA skeleton. Individual FA values were warped onto this mean skeleton mask. The resulting tract-invariant skeletons for each participant were fed into voxel-wise permutation-based cross-subject statistics. The MD, AD and RD data were also subjected to similar warping and analysis.

### 2.7 Statistical analyses

All statistical analyses were performed using commercially available software (StatSoft Statistica 12, Tulsa, OK, USA).

The clinical and demographic features of the sample were analyzed using Chi-squared, t-test, and Mann-Whitney U tests for categorical, normal continuous, and non-normal continuous variables, respectively. The genotype frequencies of SH2B3 rs3184504 were tested for Hardy-Weinberg equilibrium with a χ2 test for goodness of fit.

Given the a priori non-normal distribution of biological features, non-parametric tests were used for all the analyses. First, differences between MDD and BD patients in plasma levels of KYN, TRP, KYN/TRP ratio, and PRS for KYN were tested through non-parametric Mann-Whitney U test PRS.

#### 2.7.1 Effects of genes on circulating KYN levels and TRP breakdown ratio

Afterwards, the predictive role of PRS for KYN on KYN and KYN/TRP levels was tested in the context of generalized linear model (GLZM) with age, sex, and diagnosis as independent variables. When categorical predictors had a significant effect on the dependent variables, a separate slope regression design was performed.

Then, to test the effect of SH2B3 rs3184504 C>T SNP on PRS for KYN, plasma levels of KYN, and TRP breakdown non-parametric Kruskal-Wallis H tests were carried out; when significant effects were found, post-hoc multiple comparisons of mean ranks were performed.

#### 2.7.2 Effects of genes on WMHs and WM microstructure

Subsequently, PRS for KYN was entered into a GLZM as predictor and WMHs number and volume as dependent variables adding age, sex, and diagnosis as independent variables in each model. Similar analyses were performed considering SH2B3 rs3184504 C>T SNP as predictor.

For analyses above, when significant effects were found, post-hoc comparisons were performed through Wald statistics for parameter estimation of the factor’s different levels.

Finally, it was tested whether plasma levels of KYN and TRP breakdown could have a predictive effect on WMHs.

Afterwards, voxel-wise DTI analyses were performed using nonparametric permutation-based testing as implemented in Randomise in FSL to explore the effect of variables of interest on WM integrity. We tested for linear effect of PRS for KYN, plasma KYN, and KYN/TRP ratio on WM integrity, specifically on FA, MD, AD, and RD across the WM skeleton with general linear models (GLM). We accounted for the effects of nuisance covariates which could influence WM microstructure: age (Kochunov et al., 2007), sex (Herting et al., 2011), diagnosis (Koshiyama et al., 2020), and whether or not to had been on lithium treatment for at least 6 months at the time of the MRI (Benedetti et al., 2013). Threshold-free cluster enhancement (TFCE) was used to avoid defining arbitrary cluster-forming thresholds and smoothing levels. The data were tested against an empirical null distribution generated by 5000 permutations for each contrast, thus providing statistical maps fully corrected for multiple comparisons across space. TFCE-corrected p<0.05 were considered significant.

## 3. Results

Clinical and demographic characteristics of participants, divided according to diagnosis and genotype, are summarized in Table 1. Frequencies of SH2B3 rs3184504 C>T SNP variants were in Hardy-Weinberg equilibrium (C/C: n=28, 16%; C/T: n=96, 54.9%; T/T: n=51, 29.1%; H-W χ^2^=2.372, p=0.124). The frequency of the T allele was 56.6%.

**Table 1.** Clinical-demographic variables. Means/medians ± standard deviations/quartile range are reported. Chi-square, t-tests, and Mann-Whitney U tests refer to the comparison between the overall sample of MDD and BD.

| | MDD (n=95) | | | | BD (n=80) | | | | $\chi^2/t/z$ | p |
| --- | --- | --- | --- | --- | --- | --- | --- | --- | --- | --- |
|  | Overall | AA (n=27) | AB (n=55) | BB (n=13) | Overall | AA (n=24) | AB (n=41) | BB (n=15) |  |  |
| Age (years) | 50.705<br>±<br>10.343 | 52.889<br>±<br>9.162 | 50.236<br>±<br>10.414 | 48.154<br>±<br>12.219 | 47.963<br>±<br>11.948 | 47.917<br>±<br>10.492 | 48.268<br>±<br>13.402 | 47.200<br>±<br>10.517 | 1.628 | 0.105 |
| Sex (M/F) | 40/55 | 13/14 | 20/35 | 7/6 | 29/51 | 11/13 | 14/27 | 4/11 | 0.623 | 0.430 |
| Education (years) | 12.904<br>±<br>6.616 | 12.481<br>±<br>3.446 | 13.019<br>±<br>3.955 | 13.308<br>±<br>2.428 | 12.430<br>±<br>3.533 | 12.261<br>±<br>3.793 | 12.439<br>±<br>3.647 | 12.667<br>±<br>2.968 | 0.868 | 0.387 |
| BMI | 24.872<br>±<br>4.246 | 24.813<br>±<br>4.859 | 24.832<br>±<br>4.054 | 25.188<br>±<br>3.976 | 25.898<br>±<br>5.187 | 26.501<br>±<br>4.736 | 24.249<br>±<br>4.365 | 29.152<br>±<br>6.263 | -1.419 | 0.158 |
| Onset | 33.218<br>±<br>12.655 | 32.769<br>±<br>11.931 | 33.176<br>±<br>13.136 | 34.600<br>±<br>13.176 | 28.380<br>±<br>10.927 | 28.875<br>±<br>11.600 | 29.195<br>±<br>11.457 | 25.143<br>±<br>7.784 | 2.624 | 0.010 |
| Illness duration (years) | 17.686<br>±<br>10.971 | 19.269<br>±<br>11.362 | 17.840<br>±<br>10.931 | 12.800<br>±<br>9.682 | 19.582<br>±<br>12.028 | 19.083<br>±<br>12.079 | 19.073<br>±<br>12.809 | 21.929<br>±<br>9.895 | -1.059 | 0.291 |
| Depressive episodes (number) | 4.506<br>±<br>4.019 | 4.261<br>±<br>2.005 | 5.044<br>±<br>5.022 | 2.818<br>±<br>1.250 | 6.289<br>±<br>5.919 | 6.174 ±<br>5.424 | 6.282<br>±<br>6.291 | 6.500<br>±<br>6.048 | -1.202 | 0.029 |
| HDRS score | 21.700<br>±<br>6.140 | 23.308<br>±<br>6.263 | 20.364<br>±<br>6.411 | 23.400<br>±<br>3.507 | 20.871<br>±<br>5.207 | 21.857<br>±<br>6.466 | 21.778<br>±<br>4.722 | 17.000<br>±<br>3.795 | 0.602 | 0.549 |
| BDI score | 14.758<br>±<br>8.250 | 14.400<br>±<br>10.596 | 15.150<br>±<br>7.265 | 13.333<br>±<br>5.132 | 15.129<br>±<br>7.415 | 15.857<br>±<br>7.904 | 15.556<br>±<br>7.302 | 13.000<br>±<br>8.173 | -0.189 | 0.851 |
| KYN (µg/ml) | 0.068<br>±<br>0.061 | 0.072<br>±<br>0.065 | 0.068<br>±<br>0.058 | 0.061<br>±<br>0.074 | 0.095<br>±<br>0.070 | 0.086 ±<br>0.063 | 0.097<br>±<br>0.075 | 0.116<br>±<br>0.051 | 3.373 | <0.001 |
| TRP (µg/ml) | 11.508<br>±<br>3.673 | 11.933<br>±<br>2.318 | 11.022<br>±<br>3.982 | 10.992<br>±<br>4.706 | 11.003<br>±<br>4.174 | 11.406<br>±<br>5.098 | 10.768<br>±<br>3.673 | 11.434<br>±<br>2.537 | 0.929 | 0.334 |
| KYN/TRP*1000 | 6.281<br>±<br>5.775 | 5.366<br>±<br>6.656 | 6.492<br>±<br>4.189 | 5.652<br>±<br>5.759 | 9.598<br>±<br>5.969 | 8.861 ±<br>5.053 | 9.985<br>±<br>4.370 | 9.916<br>±<br>5.544 | 4.005 | <0.001 |
| KYN PRS | 0.142<br>±<br>0.208 | 0.204<br>±<br>0.160 | 0.123<br>±<br>0.218 | 0.058<br>±<br>0.137 | 0.169<br>±<br>0.202 | 0.216 ±<br>0.181 | 0.169<br>±<br>0.240 | 0.082<br>±<br>0.087 | -0.966 | 0.334 |
| WMHs (number) | 9.000<br>±<br>7.000 | 10.000<br>±<br>12.000 | 9.000<br>±<br>7.000 | 7.500<br>±<br>5.000 | 11.000<br>±<br>7.000 | 12.000<br>±<br>7.000 | 10.000<br>±<br>7.000 | 11.000<br>±<br>10.000 | -0.904 | 0.366 |
| WMHs (mean volume) | 0.660<br>±<br>1.323 | 0.717<br>±<br>1.714 | 0.685<br>±<br>1.523 | 0.500<br>±<br>0.728 | 0.822<br>±<br>1.933 | 0.926 ±<br>2578 | 0.747<br>±<br>1.898 | 0.713<br>±<br>1.092 | -0.883 | 0.377 |

### 3.1 Effects of genes on circulating KYN levels and TRP breakdown ratio

Patients with BD had significantly higher circulating levels of KYN than patients with MDD (Mann-Whitney U test, Z_adj_=3.373, p=0.001), but not of TRP, with a higher TRP breakdown index (Z_adj_=4.005, p<0.001). PRS for KYN were nominally higher in patients with BD, but not significantly different in the two diagnostic groups of BD and MDD (Figure 1A).

**Figure 1.**
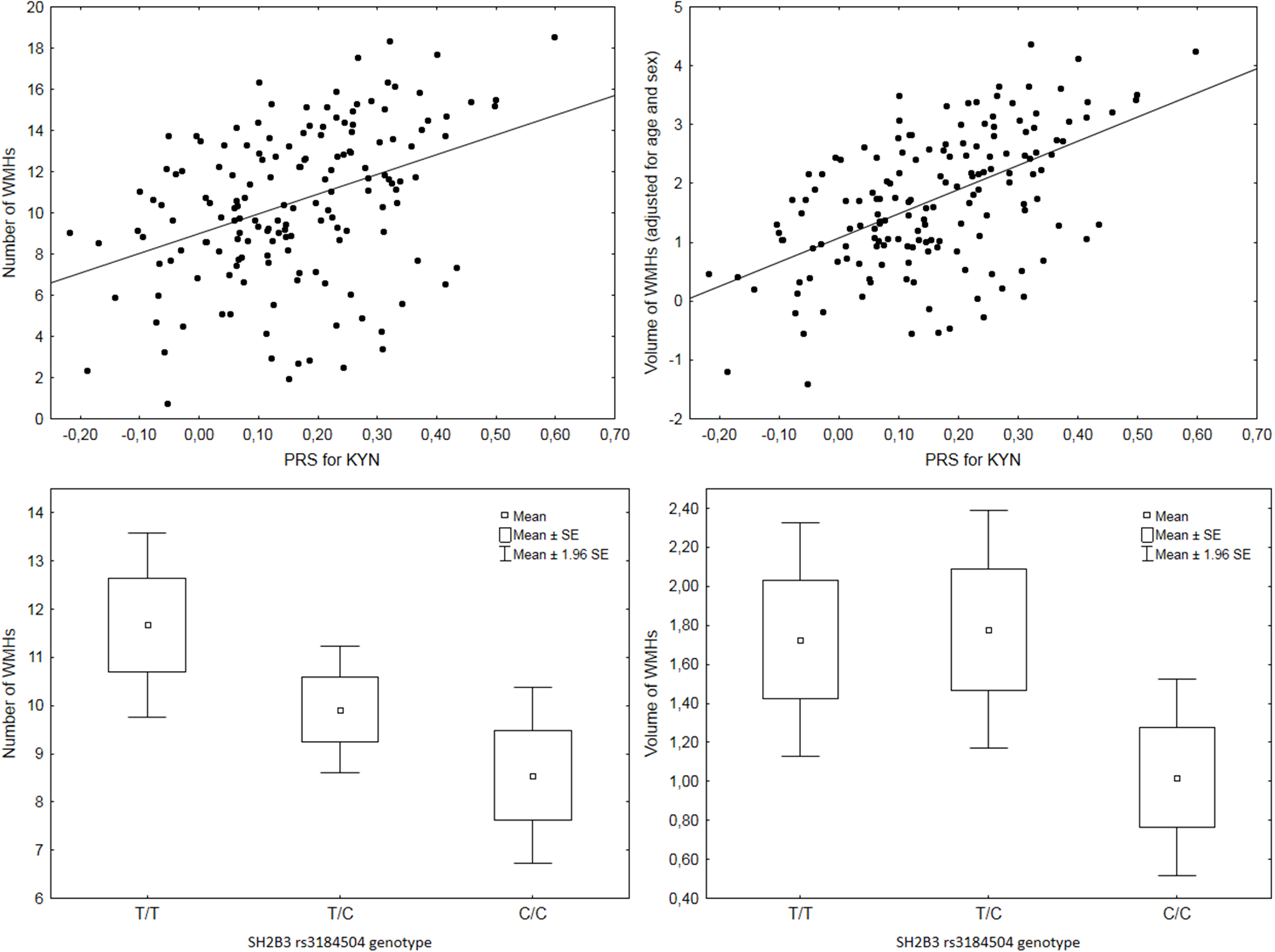
Effects of genes and diagnosis on circulating KYN levels and TRP breakdown ratio. A: Effect of diagnosis on plasma KYN levels and on PRS for KYN (means, standard errors, and confidence limits). B: Effect of PRS for KYN on circulating KYN plasma levels in patients with BD (black dots, solid line) or MDD (white dots, dotted line). D: Effect of PRS for KYN on TRP breakdown (KYN/TRP ratio) in patients with BD (black dots, solid line) or MDD (white dots, dotted line). C: Effect of SH2B3 rs3184504 genotype on PRS for KYN (means, standard errors, and confidence limits).

At GLZM regression modelling, the PRS for KYN significantly predicted circulating levels of KYN (higher PRS, higher KYN; LR χ^2^=6.172, p=0.013), with a significant contribution of diagnosis (LR χ^2^=8.038, p=0.046), but not of age and sex. Again, a separate-slopes regression showed a significant interaction of PRS and diagnosis (LR χ^2^=13.749, p=0.001), due to a higher effect of PRS in BD than in MDD (Figure 1B). Moreover, the PRS for KYN significantly predicted the TRP breakdown ratio (LR χ^2^=6.215, p=0.003), with a significant contribution of diagnosis (LR χ^2^=9.463, p=0.002), but not of age and sex. Again, a significantly higher effect of PRS in BD than in MDD was detected at separate-slopes regression (LR χ^2^=13.271, p=0.001) (Figure 1D).

The SH2B3 rs3184504 C>T SNP significantly influenced the PRS for KYN (Kruskal-Wallis ANOVA: H=14.345, p =0.001) with homozygotes for the missense rs3184504*T variant showing higher scores than rs3184504*C homozygotes (multiple comparisons z’=3.765, p=0.001), and T/C heterozygotes showing intermediate values (Figure 1D). However, rs3184504 did not significantly influence neither plasma KYN levels, nor TRP/KYN ratio.

### 3.2 Effects of genes on WMHs and WM microstructure

At GLZM regression modelling, the PRS for KYN significantly predicted the number of WMHs (higher PRS, more WMHs; LR χ^2^=6.172, p=0.013), with a major contribution of age (LR χ^2^=52.790, p<0.001), but not of diagnosis and sex. The PRS for KYN also predicted the mean volume of WMHs (higher PRS, bigger WMHs; LR χ^2^=9.587, p=0.002), again with a major contribution of age (LR χ^2^=19.415, p<0.001), but not of diagnosis and sex (Figure 2, top).

**Figure 2.**
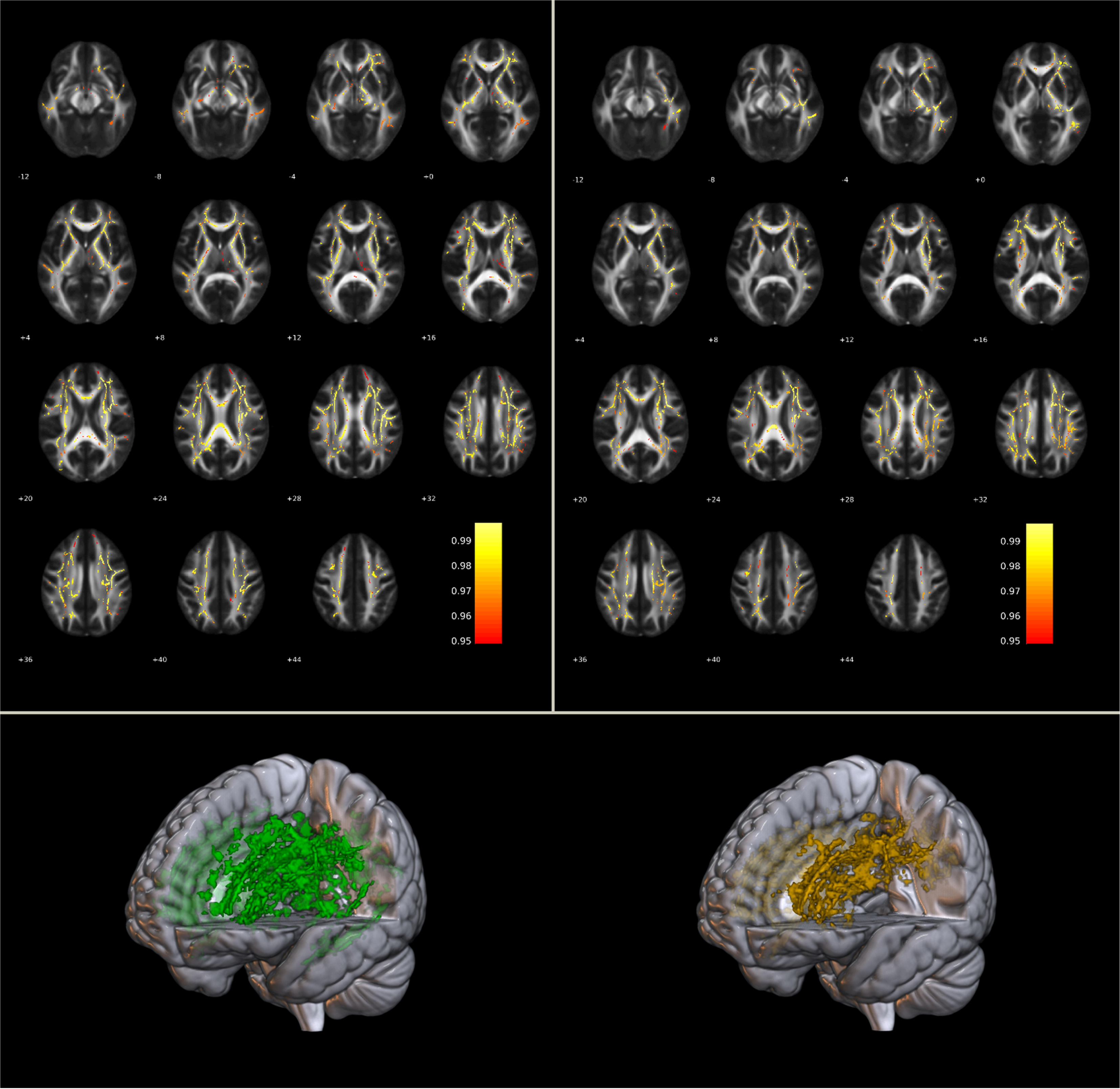
Effects of genes on WMHs. Top: effects of PRS for KYN on number and total volume of WMHs, adjusted for age and sex. Bottom: effect of SH2B3 rs3184504 genotype on number and total volume of WMHs (means, standard errors, and confidence limits).

The SH2B3 rs3184504 C>T SNP significantly influenced the number of WMHs (LR χ^2^=7.795, p=0.020) with homozygotes for the missense rs3184504*T variant showing higher scores than rs3184504*C homozygotes (multiple comparisons parameter estimate: b=1.75, W2=7.614, p=0.006), and T/C heterozygotes showing intermediate values; again with a major contribution of age (LR χ^2^=52.479, p<0.001), but not of diagnosis and sex. Inspection of data shows that homozygous and heterozygous carriers of the rs3184504*T variant show nominally bigger mean WHMs volumes than C/C homozygotes, but the effect was not significant (Figure 2, bottom).

Levels of circulating KYN and of KYN/TRP ratio did not significantly influence WMHs number and volume.

Dealing with TBSS analyses, KYN PRS showed to have a significant positive effect on L1 (p=0.039; Figure 3, left), MD (p=0.020; Figure 3, right), and a trend towards significance for RD (p=0.098) (Table 2).

**Figure 3.**
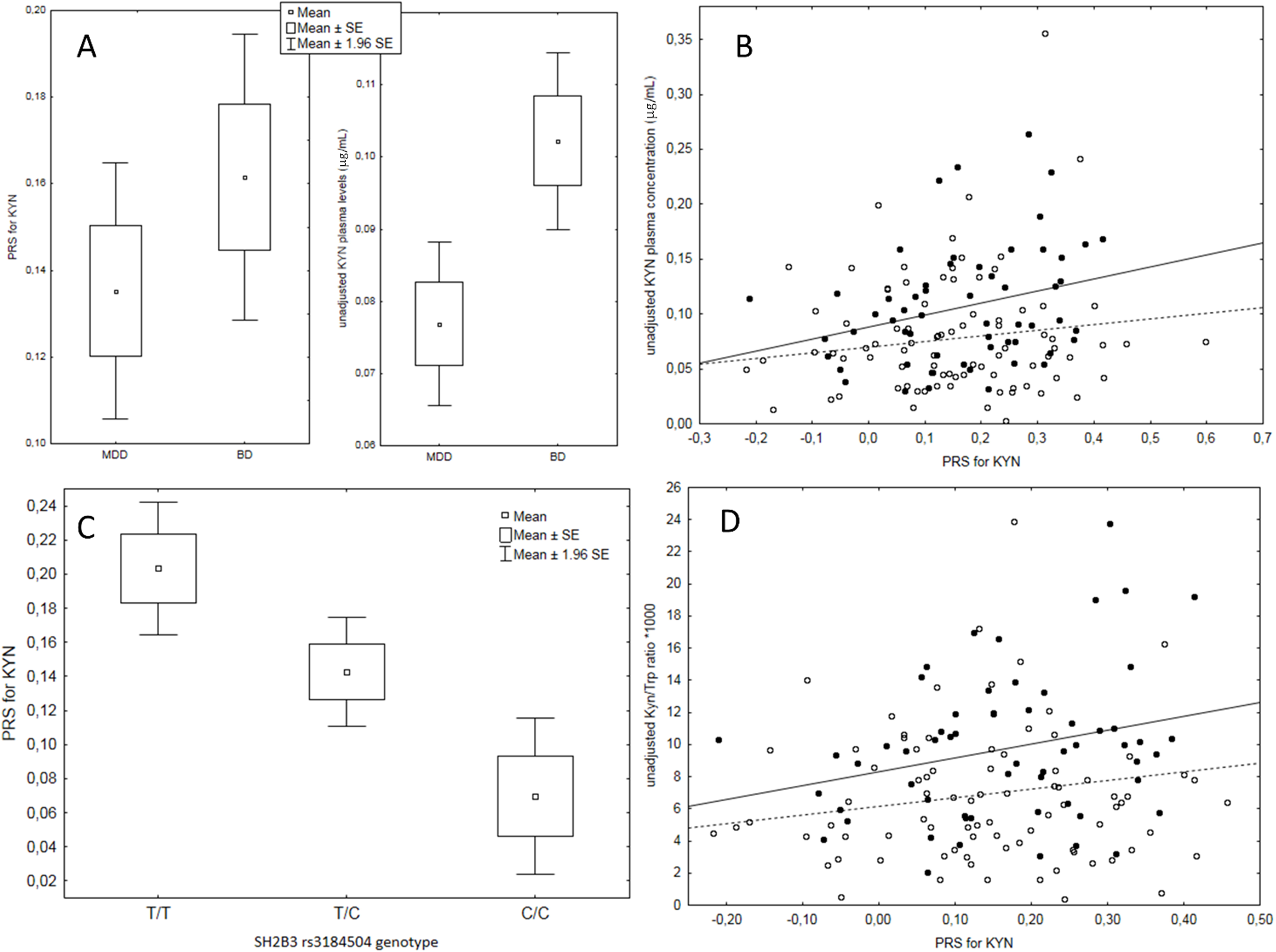
Effect of PRS for KYN on WM microstructure. On the left WM tracts positively associated with AD; on the right WM tracts positively associated with MD. Bottom: in green and orange the 3D rendering of WM tracts associated with AD (left) and MD (right), respectively.

**Table 2.** Table reporting TBSS analyses results. Mean ± standard deviation of L1 and MD, cluster characteristics, and involved tracts are reported.

| DTI Measure | No. voxels & signal peaks (x, y, z) | White matter tracts |
| --- | --- | --- |
| L1<br>0.122 ± 0.003 | 32259<br>34; 6; 20 (R Superior longitudinal fasciculus) | Anterior thalamic radiation L<br>Anterior thalamic radiation R<br>Corticospinal tract L<br>Corticospinal tract R<br>Cingulum (cingulate gyrus) L<br>Cingulum (cingulate gyrus) R<br>Forceps major<br>Forceps minor<br>Inferior fronto-occipital fasciculus L<br>Inferior fronto-occipital fasciculus R<br>Inferior longitudinal fasciculus L<br>Inferior longitudinal fasciculus R<br>Superior longitudinal fasciculus L<br>Superior longitudinal fasciculus R<br>Uncinate fasciculus L<br>Uncinate fasciculus R |
| MD<br>0.076 ± 0.003 | 25079<br>41; 6; 18 (R Superior longitudinal fasciculus) | Anterior thalamic radiation L<br>Anterior thalamic radiation R<br>Corticospinal tract L<br>Corticospinal tract R<br>Cingulum (cingulate gyrus) L<br>Cingulum (cingulate gyrus) R<br>Forceps major<br>Forceps minor<br>Inferior fronto-occipital fasciculus L<br>Inferior fronto-occipital fasciculus R<br>Inferior longitudinal fasciculus L<br>Inferior longitudinal fasciculus R<br>Superior longitudinal fasciculus L<br>Superior longitudinal fasciculus R<br>Uncinate fasciculus L<br>Uncinate fasciculus R |

We found no significant effects when considering plasma levels of KYN and KYN/TRP breakdown.

## 4. Discussion

The main findings of the present study are that a PRS for KYN, as estimated in the general population, also predicts circulating levels of KYN and TRP breakdown in patients affected by mood disorders; that the same PRS contributes to an enhanced neurovascular risk, proportionally increasing number and volumes of WMHs; and that the SH2B3 rs3184504 C>T SNP is genetically associated with the PRS for KYN, but not with circulating KYN levels, and influences WMHs of presumed vascular origin.

In agreement with previous studies by our group, we observed higher levels of KYN and TRP breakdown in patients with BD than in patients with MDD. Given that patients with BD show higher circulating levels of pro-inflammatory cytokines, which activate IDO (Poletti et al., 2021), this finding is likely to reflect a greater activation of inflammatory mechanism in this diagnostic group (Poletti et al., 2024).

The abnormally high activation of IDO triggers a cascade of events in patients with mood disorders (see Introduction), including effects on serotoninergic and glutamatergic neurotransmission, that, when modelled in rodents, lead to the development of depressive-like behaviors, and could play a major role in the pathophysiology of depressive symptoms in humans (Pocivavsek et al., 2024). Possibly because of the perturbation of core neurotransmitters which influence brain homeostasis at multiple levels, including synaptic neuroplasticity and astrocyte and oligodendrocyte cell functions, circulating levels of KYN pathway metabolites associate with detrimental effects on brain grey and white matter in patients with mood disorders. Genetic factors affecting IDO activation could then influence multiple phenotypic traits, to predict clinical outcomes of the disorders, including neuroprogression and the risk of neurovascular insults.

In particular, higher individual scores at the PRS for KYN associated with both, higher KYN and TRP breakdown levels, and with an increasing number and volume of WMHs. However, neither KYN, nor TRP breakdown, associated with these brain signs of microvascular pathology. KYN causes endothelium-independent vasodilation via K+ channels in the vascular smooth muscle (Sakakibara et al., 2015; Worton et al., 2021), and it could possibly exert an influence in regulating vascular tone and hemodynamic responses in human physiology, as suggested by the observation that arteries of normotensive and hypertensive pregnant women are similarly responsive to vasodilating KYN (Worton et al., 2021). The possible role of KYN in cardiovascular disease is highly complex, and it has been suggested that metabolites within the KYN pathway could have cardiovascular protective effects (Yang et al., 2024). Moreover, KYN crosses the blood-brain barrier, and in animal models the peripheral administration of KYN increases the cerebral blood flow, both in normal and in ischemic conditions (Sas et al., 2003).

However, the PRS for KYN showed a strong association with number and volume of WMHs, as did the SH2B3 rs3184504 genotype. In turn, rs3184504 markedly influenced individual PRS for KYN scores, but not circulating levels of KYN. In agreement with previous studies, this might suggest that the genetic landscape for KYN includes gene variants that are closely related to inflammation, but not directly related to KYN levels (Bagheri et al., 2021). SH2B3 plays a role in protection against infection, with the mutant rs3184504*T allele promoting a stronger immune response and a better survival to sepsis (Allenspach et al., 2021), and in the early stages of sepsis the induction of IDO first elicits strong proinflammatory effects, with KYN thereafter contributing to suppress T-cell response, and to maintain immunological tolerance by innate immunity cells (Gaelings et al., 2017; Krupa et al., 2022).

In this respect, previous research by our group showed that circulating immune cell composition and activation status associate with WM integrity in mood disorders, with different effects of Th17, NK, and CD8+ T cells (Poletti et al., 2024). In particular, activated CD8+ T cell populations negatively associated with WM microstructure (Aggio et al., 2023), and the rs3184504*T allele associates with enhanced CD8+ T cells effector functions. It is then tempting to speculate that the same selective pressure to promote survival against infective pathogens by enhancing immediate, inflammatory responses, could have jointly influenced these genetic factors, to allow for rapid adaptation in similar environmental conditions.

In line with these results, the PRS for KYN, found to be influenced by rs3184504 SNP, was positively associated with AD and MD at DTI, with a trend towards significance for RD in our sample. The widespread pattern of tracts significantly affected overlaps considering the two different DTI measures encompassing corpus callosum, bilateral cingulum bundle, inferior fronto-occipital fasciculus, inferior and superior longitudinal fasciculi, and uncinate fasciculus.

Notably, together with the occurrence of WMHs, WM microstructure disruption is a neurobiological biomarker of both mood disorders and cardiovascular diseases (Favre et al., 2019; Li et al., 2024; Moroni et al., 2018; Silva et al., 2024; Thomas et al., 2002; van Velzen et al., 2020). In particular, our results showing PRS associated with higher AD and MD, along with a trend in the same direction for RD, speak of axonal integrity alterations, and myelin disruption with possible vasogenic edema, conditions already observed in patients with cardiovascular and cardiometabolic diseases (Kumar et al., 2011; McBride et al., 2022; Moroni et al., 2018), and cognitive performance (Power et al., 2019; Wang et al., 2022; Zanon Zotin et al., 2022).

More specifically, extracellular edema occurs as a typical reaction to inflammation with alterations in BBB, allowing plasma-like fluid to leak from blood vessels into the extracellular space, with water molecules diffusing more freely in all directions, as reflected by higher MD, AD, and RD (Assaf and Pasternak, 2008; Aung et al., 2013). Interestingly, aquaporin 4 (AQP4), a specialised water channel that regulates the water flow across the BBB, was found to be positively associated with the rate of IDO and TDO activation (Du et al., 2020), possibly promoting edema formation (Ito et al., 2005). Coherently, dexamethasone administration with consequent downregulation of TDO expression is used as a medication for the mitigation of brain edema (Kostaras et al., 2014; Shannon et al., 2015).

Given previous results linking Kyn PRS to WMHs, it is likely that a higher genetic predisposition to inflammatory, vascular, and endothelial alterations could lead to neurovascular disruptions, which affect cerebral blood flow and WM structure.

Notably, the pattern of our results overlaps with the widespread pattern of WM tracts affected by inflammatory compounds in MDD and BD (Benedetti et al., 2016; Lim et al., 2021). Since we speculated about the immune-inflammatory underground of Kyn PRS, this could foster immune dysregulation in depression, thus affecting WM microstructure.

Finally, our data warrant interest in further study of the genetic landscape of KYN in clinical samples. The association of the PRS and KYN circulating levels in our clinical sample suggests that the core elements of this PRS are shared with the general population (Canadian participants), where it has been estimated (Chen et al., 2023). PRSs may include high levels of variance in genetic architecture among the polymorphisms in population-specific PRSs for the same trait (Graham et al., 2021), and further research is then needed to explore if patients with mood disorders might show differences in respect to healthy participants. Moreover, a comparison with published studies in the Italian population shows that rs3184504*T allele frequencies in our patients with mood disorders were significantly higher than those expected in control participants (0.566 vs 0.494, χ^2^=4.44, p=0.0351) (Zhernakova et al., 2010), and similar to those observed in Italian clinical samples with inflammatory conditions, such as coeliac disease (0.554) (Romanos et al., 2009). SH2B3 rs3184504 shows a high variance in allele frequency among populations and a selective sweep, with the rs3184504*T allele being uniquely enriched in samples of European descent (Zhernakova et al., 2010). It is tempting to surmise that SH2B3 gene variants could influence the risk for both inflammatory conditions and mood disorders. Further research is needed to test this hypothesis in carefully selected samples because the variance of rs3184504 allele frequency is also high among European groups.

While this study provides valuable insights into the genetic landscape of Kyn in patients with mood disorders, some limitations should be acknowledged. The patients included in our study were not drug-naive, which may introduce some bias induced by the effects that lifetime treatments can have on most of the measures used in this work. Another limitation is the lack of ethnic variability within our sample, which restricts the generalizability of our findings across different populations. This latter limitation couples with the single-centre nature of the present study, restricting the possibility of reproducing our results in different samples. Finally, possible interactions with other metabolic or inflammatory PRS could modulate the observed associations.

## Conclusions

The present study highlights the relevance of focusing on the KYN pathway as a possible common genetic underground between cardiometabolic disease and mood disorders, pointing to immune-inflammatory alterations. In particular, the genetic influence on KYN was shown to be strictly dependent on the rs3184504 polymorphism, affecting outcomes of presumed vascular suffering, such as WMHs, and WM microstructure. These findings offer valuable insights into the pathophysiological mechanisms underlying mood disorders and may guide further investigation of these biosignatures, ultimately integrating them into precision psychiatry approaches.

## Data Availability

All data produced in the present study are available upon reasonable request to the authors

## Conflict of interest

None

## Acknowledgment

Funded by the Italian Ministry of Health, grant PNRR-MAD-2022-12375859 Inflammation and depression: a multidisciplinary approach to dissect pathogenetic mechanisms in the onset, comorbidity and treatment response.

## Notes

### Competing Interest Statement

The authors have declared no competing interest.

### Funding Statement

Funding for this study was provided by the Italian Ministry of Health, Grant PNRR-MAD-2022-12375859

### Author Declarations

The local ethical committee approved all the research activities (IRCCS Ospedale San Raffaele) and complied with the Helsinki Declaration of 1975, as revised in 2013.

